# Vagus Nerve Stimulation in Failed Epilepsy Surgery: 36 Month Outcomes From the CORE-VNS Study

**DOI:** 10.64898/2026.04.17.26351099

**Authors:** Evan N. Nicolai, Katarzyna Sieradzan, Olaf Schijns, Menaka P. Fry, Kim Rijkers, Ryan Verner, Saleh Baeesa, Nilesh Kurwale, Gaia Giannicola, Charles Gordon, Alex Moon, Francesca Beraldi, Arjune Sen, David A. Mays

**Affiliations:** LivaNova PLC (or a subsidiary), London, England, United Kingdom; Southmead Hospital (North Bristol NHS Trust), Bristol, England, United Kingdom; Department of Neurosurgery, Academic Center for Epileptology, Maastricht University Medical Center, The Netherlands; Mental Health and Neuroscience Research Institute (MHeNs), Maastricht University, Maastricht, The Netherlands; Department of Neurosurgery, Cleveland Clinic, Ohio; Neurosciences Department, King Faisal Specialist Hospital and Research Center, Jeddah, Saudi Arabia; Deenanath Mangeshkar Hospital and Research Center, Pune, India; Valos S.r.L, Genova, Italy; Oxford Epilepsy Research Group, Nuttfield Department of Clinical Neurosciences, John Radcliffe Hospital, Oxford, UK

**Keywords:** Brain Surgery, Drug-resistant, Epilepsy, Epilepsy Surgery, Neuromodulation, Vagus nerve stimulation

## Abstract

**Objective:** Vagus nerve stimulation (VNS) is an established neuromodulation therapy used in the management of drug-resistant epilepsy (DRE), or when other intracranial surgical modalities have not reduced seizure burden. We evaluated whether prior intracranial surgery for epilepsy influences safety and effectiveness outcomes with adjunctive VNS, using real-world data from the CORE-VNS study.

**Methods:** CORE-VNS (NCT03529045), a prospective, multicenter, international observational study, was designed to collect data on seizure and non-seizure outcomes in patients with DRE treated with VNS. Participants were identified as having or not having undergone prior intracranial brain surgery for epilepsy (ICSE) and received an initial VNS implant. Baseline seizure frequency data and patient-reported outcome measures were collected at 3, 6, 12, 24, and 36 months. This analysis compared the baseline data for VNS therapy and safety outcomes at 36 months.

**Results:** Among 531 participants implanted with VNS, prior ICSE was performed in 84. Median percentage seizure reductions at 36 months for all seizures (76.6% and 76.3%), all focal seizures (83.3% and 71.8%), and all generalized seizures (77.8% and 76.2%) were found to be similar between those without and with a history of ICSE, respectively. The 50% responder rate for all seizures reported at baseline was similar, 64.8% and 61.8%, in both groups and complete seizure freedom was reported by 17.9% and 8.8%, respectively. Implant-related adverse events (AE) and serious AE rates were similar between groups.

**Conclusion:** VNS was associated with clinically meaningful seizure reductions and showed a consistent safety profile irrespective of the history of ICSE. Prior ICSE should not be a contraindication to the consideration of VNS.

## Introduction

Seizures persist in a subgroup of people with epilepsy (PwE) following multiple trials of antiseizure medications (ASMs) and, for some, despite intracranial surgery for epilepsy (ICSE).^1^ Even after neurosurgical intervention, seizure freedom (SF) rates vary, and a considerable subset continues to experience disabling seizures requiring ongoing pharmacotherapy.^1,2^ Vagus nerve stimulation (VNS) is an established neuromodulation therapy for drug-resistant epilepsy (DRE) and has demonstrated broad effectiveness across seizure types in real-world settings.^3,4^ However, it remains unclear whether ICSE influences outcomes with subsequent VNS. This question is of high clinical relevance when considering the sequencing of therapies.

During the clinical trials that led to FDA approval for VNS Therapy in DRE, ICSE was not common enough to allow for adequate analysis of possible effects on outcomes. At the time of those early clinical studies, the mechanism for VNS was not well understood and there was reasonable concern that previous brain surgery may affect VNS action. Consequently, the current VNS Therapy label reads “Safety and Efficacy Not Established” in patients with a history of prior brain surgery.

In the 30 years since VNS was approved for use in DRE, understanding of its mechanisms of action has evolved. Briefly, the sensory afferents within the vagus nerve project to the nucleus tractus solitarius (NTS) in the brainstem, which in turn projects to locus coeruleus (LC) and raphe nucleus (RN), which provide broad projections of noradrenergic and serotonergic signaling throughout the brain.^5,6^ Intracranial surgical options for epilepsy include resection of the epileptogenic zone (EZ), (hemi-) cortical disconnection procedures, and intracranial neuromodulation. Here, we evaluate the effectiveness and safety of VNS Therapy in PwE who did not become seizure-free after ICSE.

Several studies of VNS Therapy have been conducted in PwE who have had a positive history of ICSE. In these cases, VNS was used on a compassionate-use basis; however, the number of subjects has been insufficient to meaningfully assess the impact of prior surgery on outcomes.^7,8^

We leveraged the large, multinational CORE-VNS dataset to compare safety and seizure-frequency outcomes in PwE and DRE based on whether or not they had previous ICSE.^4,9^ Our hypotheses were 1) that the effectiveness of VNS would not be different in people who had undergone prior ICSE since broad activation occurs via brainstem structures first before spreading to cortex, ^5,6^and 2) that the adverse effects – such as cough or voice alteration – of VNS would not be changed since these are due to activation of the motor efferent nerve fibers of the vagus nerve traveling to the laryngeal muscles, which all happens extracranially.^10,11^

## Methods

### Study design and participant selection

CORE-VNS is an international, multicenter, prospective, observational study designed to evaluate real-world outcomes of contemporary VNS Therapy in PwE and DRE under routine clinical management.^3,4,8^ Baseline seizure frequency was reported for each participant prior to device implantation. Visits were scheduled at 3, 6, 12, 24, and 36 months post-implantation. Similar to clinical practice, seizure frequency by seizure type was collected at each visit for the preceding 3-month period. VNS-related adverse events were recorded based upon patient or caregiver report.

Two DRE subgroups were defined at baseline: PwE with a positive history of ICSE and PwE without ICSE (Table 1). No formal power calculation was conducted because this was a post-hoc subset analysis within an open-label observational study. The study was not prospectively designed to detect small differences. Between-group comparisons were performed to assess whether outcomes were broadly comparable across subgroups.

**Table 1.** Demographics.

|  | PRIOR INTRACRANIAL BRAIN<br>SURGERY FOR<br>EPILEPSY<br>N=84 | NO PRIOR INTRACRANIAL<br>BRAIN SURGERY FOR<br>EPILEPSY<br>N=447 |
| --- | --- | --- |
| **AGE AT INFORMED CONSENT (YEARS)** |  |  |
| MEAN (SD) | 25.5 (14.6) | 23.5 (16.9) |
| MEDIAN | 23 | 19 |
| MIN, MAX | 3, 63 | 1, 71 |
| 31 DAYS-2 YEARS | - | 11 (2.5%) |
| 3-12 YEARS, N (%) | 16 (19%) | 139 (31.1%) |
| 13-17 YEARS, N (%) | 15 (17.9%) | 60 (13.4%) |
| ≥ 18 YEARS, N (%) | 53 (63.1) | 237 (53.0) |
| **SEX** |  |  |
| MALE, N (%) | 44 (52.4) | 228 (51.0) |
| FEMALE, N (%) | 40 (47.6) | 219 (49.0) |
| **NUMBER OF ASMS FAILED** |  |  |
| MEAN (SD) | 7.2 (3.0) | 6.7 (3.1) |
| MEDIAN | 7 | 6 |
| MIN, MAX | 2, 17 | 2, 20 |
| **AGE AT EPILEPSY DIAGNOSIS** |  |  |
| <4 YEARS, N (%) | 36 (42.9) | 183 (40.9) |
| 4-11 YEARS, N (%) | 23 (27.4) | 121 (27.1) |
| 12-<18 YEARS, N (%) | 9 (10.7) | 68 (15.2) |
| ≥18 YEARS, N (%) | 16 (19.0) | 75 (16.8) |
| **COGNITIVE STATUS** |  |  |
| NORMAL, N (%) | 26 (31.0) | 142 (31.8) |
| MINIMAL IMPAIRMENT, N (%) | 14 (16.7) | 97 (21.7) |
| MODERATE IMPAIRMENT, N (%) | 20 (23.8) | 114 (25.5) |
| SEVERE, N (%) | 24 (28.6) | 94 (21.0) |
| **SEIZURE TYPE AT BASELINE** |  |  |
| COMBINED, N (%) | 21 (25.0) | 170 (38.0) |
| FOCAL, N (%) | 55 (65.5) | 192 (43.0) |
| GENERALIZED, N (%) | 7 (8.3) | 77 (17.2) |
| UNKNOWN, N (%) | 1 (1.2) | 8 (1.8) |
| **MODEL OF PULSE GENERATOR** |  |  |
| SENTIVA<br>(MODEL 1000), N (%) | 42 (50.0) | 200 (44.7) |
| ASPIRESR<br>(MODEL 106), N (%) | 37 (44.0) | 202 (45.2) |
| DEMIPULSE GENERATOR (MODEL 106),<br>N (%) |  | 8 (1.8) |
| PULSE GENERATOR<br>(MODEL 02), N (%) | 5 (6.0) | 35 (7.8) |

### Outcomes

Seizure frequency was recorded at each clinic visit based on participant-reported occurrences of each seizure type during the preceding 3-month interval. The monthly seizure frequency during this period was compared with the baseline values for the same seizure types to determine the percentage change. Seizure freedom was defined as the complete absence of seizures during the reporting interval (3 months prior) to the current visit. Participants with no baseline seizures in the baseline period or no follow-up efficacy assessments were excluded from seizure reduction analyses. Responder rates, defined as the proportion of participants achieving 50%, 80%, or 100% seizure reduction, were calculated for each group.

Adverse events (AEs) were defined as any untoward medical occurrence following VNS implantation, including events that were related to the implant/procedure/stimulation/device. Serious AEs (SAEs) were classified as life-threatening events, hospitalizations, and persistent disabilities. All AEs and SAEs were collected prospectively according to the protocol and categorized by their relationship with VNS Therapy over 36 months.

## Statistical analysis

Descriptive statistics were performed to summarize results in participants with and without prior brain surgery. Continuous variables are presented as mean ± SD or median (IQR), as appropriate, and categorical variables as counts and percentages. The Wilcoxon rank-sum test was used to compare the prior ICSE and no prior ICSE subgroups for seizure reduction outcomes.

### Incidence Rate Ratio Estimation

Incidence rate ratios (IRRs) were calculated to compare adverse and serious adverse event rates between participants with prior brain surgery (N = 84) and those without prior brain surgery (N = 447). For each category, incidence rates (IRs) were computed as the number of events divided by the total exposure time within the subgroup. The total exposure time was defined as the sum of the exposure times of all participants in the subgroup, where each participant’s exposure time was calculated as the time (in years) from device implantation until the last post-implant assessment, or, for participants who withdrew from the study until the discontinuation date. The IRR was defined as the ratio of the incidence rate in the prior surgery group to that in the no prior surgery group. Ninety-five percent confidence intervals (CIs) for IRRs were estimated using a Poisson approximation, applying Equation 1 where a and b represent the event counts in each group.

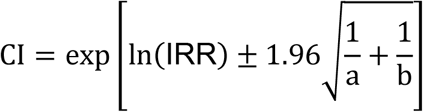

Equation 1: Used to estimate the 95% confidence interval for IRR.

This approach assumes approximate normality of the log (IRR) and independence of events across groups. When zero events were observed in a group, a continuity correction of 0.5 events was applied to estimate the incidence rate ratio and its confidence interval.

## Results

### Demographics

Among those with a history of ICSE (n = 84), 52.4% of participants were male, with a mean age of 25.5 years (SD 14.6); most were adults (≥18 years, 63.1%), with smaller proportions aged 13–17 years (17.9%) and 3–12 years (19.0%). In the group without a history of ICSE (n = 447), 51.0% were male, the mean age was 23.5 years (SD 16.9), with 53.0% adults, 13.4% adolescents, 31.1% children aged 3–12 years, and 2.5% <3 years. Participants in both cohorts were predominantly White, with a smaller proportion of Asian individuals, while other racial groups were rare.

In those who had undergone previous ICSE, lesionectomy was the most common surgery type (36.9%), followed by lobar resection (22.6%) and callosotomy (19.0%). Other surgery types accounted for less than 10% of the group and are summarized in Table 2.

**Table 2.** Previous Brain or Epilepsy Surgery (Excluding VNS)

| *Previous Brain or Epilepsy Surgery Excluding VNS* | *Prior Brain Surgery (N=84)* |
| --- | --- |
| *Lesionectomy, n (%)* | 31 (36.9%) |
| *Lobar resection* | 19 (22.6%) |
| *Callosotomy* | 16 (19.0%) |
| *Anterior temporal lobectomy* | 6 (7.1%) |
| *Anterior temporal lobectomy plus amygdalohippocampectomy* | 5 (6.0%) |
| *Lobar disconnection* | 4 (4.8%) |
| *Laser ablation* | 4 (4.8%) |
| *Multilobar resection* | 3 (3.6%) |
| *Multiple subpial transections* | 3 (3.6%) |
| *Hemispheric disconnection (hemispherotomy)* | 1 (1.2%) |
| *Intracranial Neuromodulation* | 1 (1.2%) |
| *Multilobar disconnection* | 1 (1.2%) |

### Seizure outcomes

Seizure outcomes were reported separately for participants with (n = 84) and without (n = 411) ICSE, excluding 37 implanted participants who had either zero recorded seizures at baseline, were missing baseline seizure assessment, or had no follow-up assessments (Table 3). For people without ICSE, median reductions from baseline in seizure frequency ranged from 44.4% (95% CI = 33.33 to 50.00) at 3 months to 76.6% (95% CI = 66.67 to 86.36) at 36 months for all seizures (Figure 1). For people with a history of ICSE, reductions were similar in direction and magnitude, ranging from 37.9% (95% CI = 10.91 to 53.85) at 3 months to 76.3% (95% CI = 48.57 to 87.50) at 36 months.

**Table 3.** Monthly Seizure Statistics.

| Month | Prior Brain Surgery |  |  | No Prior Brain Surgery |  |  |
| --- | --- | --- | --- | --- | --- | --- |
|  | *n* | Mean<br>(SD) | Median<br>(CI) | *n* | Mean<br>(SD) | Median<br>(CI) |
| **All Seizures** |  |  |  |  |  |  |
| *Baseline* | 83 | 71.27<br>(147.13) | 24.00<br>(14.33, 36.67) | 411 | 75.62<br>(263.20) | 15<br>(10.67, 20.00) |
| 3 | 72 | 47.43<br>(115.38) | 13.67<br>(5.33, 20.00) | 330 | 43.01<br>(193.43) | 7.17<br>(5.00, 10.00) |
| 6 | 71 | 55.23<br>(152.17) | 9.67<br>(4.00, 21.33) | 330 | 46.16<br>(172.01) | 6.00<br>(4.00, 10.00) |
| 12 | 72 | 33.07<br>(58.70) | 7.50<br>(4.67, 14.33) | 358 | 43.59<br>(197.20) | 4.67<br>(3.67, 7.00) |
| 24 | 70 | 40.92<br>(116.34) | 10.83<br>(3.0, 17.33) | 337 | 27.31<br>(81.27) | 3.33<br>(2.00, 5.00) |
| 36 | 68 | 25.62<br>(44.11) | 6.00<br>(2.00, 20.00) | 324 | 20.64<br>(43.67) | 3.00<br>(2.00, 4.33) |
| **Focal Overall** |  |  |  |  |  |  |
| *Baseline* | 74 | 68.79<br>(154.28) | 20.00<br>(10.00, 30.33) | 314 | 55.08<br>(205.96) | 13.33<br>(10.00, 20.00) |
| 3 | 67 | 46.97<br>(119.46) | 10.00<br>(4.33, 16.67) | 254 | 37.09<br>(214.45) | 5.17<br>(3.33, 8.00) |
| 6 | 64 | 55.93<br>(160.65) | 8.00<br>(3.33, 17.00) | 251 | 38.24<br>(134.85) | 5.00<br>(3.33, 9.67) |
| 12 | 64 | 30.83<br>(59.73) | 6.33<br>(4.33, 13.00) | 269 | 41.54<br>(219.14) | 4.00<br>(2.33, 6.33) |
| 24 | 63 | 39.85<br>(121.47) | 9.33<br>(2.00, 16.00) | 253 | 26.43<br>(88.50) | 2.67<br>(1.67, 4.33) |
| 36 | 60 | 24.72<br>(44.96) | 4.67<br>(2.00, 13.33) | 249 | 16.79<br>(40.58) | 2.33<br>(1.33, 3.67) |
| **Generalized Overall** |  |  |  |  |  |  |
| *Baseline* | 13 | 50.64<br>(57.08) | 33.33<br>(1.00, 75.00) | 152 | 77.22<br>(301.54) | 8<br>(6.00, 14.67) |
| 3 | 9 | 23.59<br>(33.31) | 10.00<br>(0.00, 44.00) | 120 | 34.84<br>(65.30) | 4.17<br>(2.00, 9.33) |
| 6 | 10 | 29.70<br>(34.79) | 14.50<br>(0.33, 63.33) | 119 | 28.54<br>(55.59) | 4.00<br>(1.67, 9.00) |
| 12 | 10 | 25.83<br>(30.97) | 19.83<br>(0.33, 45.33) | 131 | 30.32<br>(67.55) | 4.00<br>(1.00, 6.67) |
| 24 | 10 | 30.87<br>(52.29) | 2.50<br>(0.00, 100.00) | 124 | 19.29<br>(42.18) | 2.00<br>(0.67, 4.00) |
| 36 | 11 | 20.82<br>(36.17) | 5.33<br>(1.00, 50.00) | 115 | 20.84<br>(41.36) | 1.67<br>(0.67, 4.00) |

**Figure 1.**
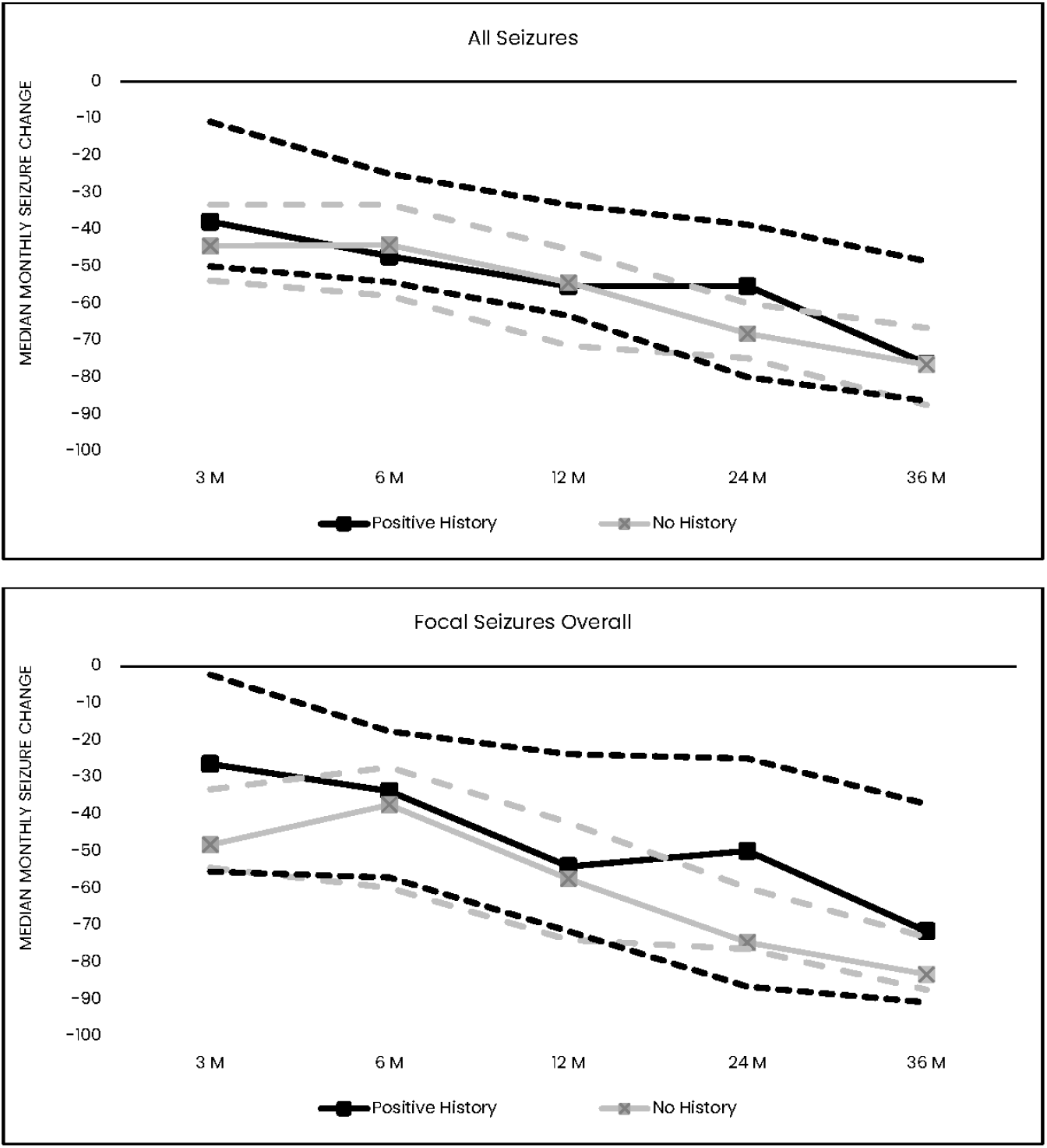
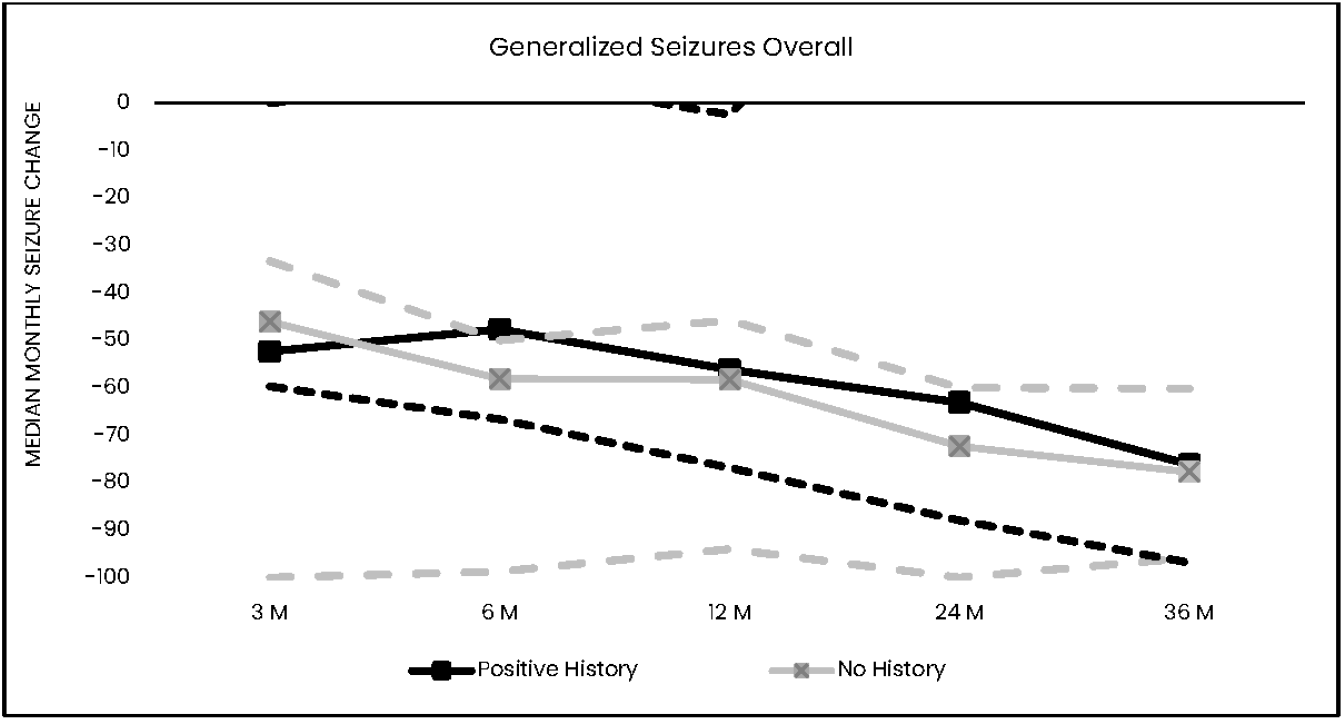
Median Monthly Seizure Frequency Change and Confidence Intervals by Study Visit. Solid lines represented median change from baseline seizures. Dashed lines represent median 95% Cl for each identical colored group. Values non-significant at each time point, Wilcoxon rank-sum test comparing prior surgery vs no prior surgery subgroups at each visit.

At 36 months, 64.8% of those without a history of ICSE had a ≥ 50%, 47.8% had ≥ 80%, and 29.9% had a 100% seizure reduction from baseline. Similarly, 61.8% of participants with a history of ICSE achieved ≥ 50%, 47.1% achieved ≥ 80%, and 16.2% achieved 100% seizure reduction from all seizures reported at baseline. Complete seizure freedom from all seizures, including those not reported at baseline, was noted in 17.9% of participants without a history and 8.8% of those with a history of ICSE (Figure 2).

**Figure 2.**
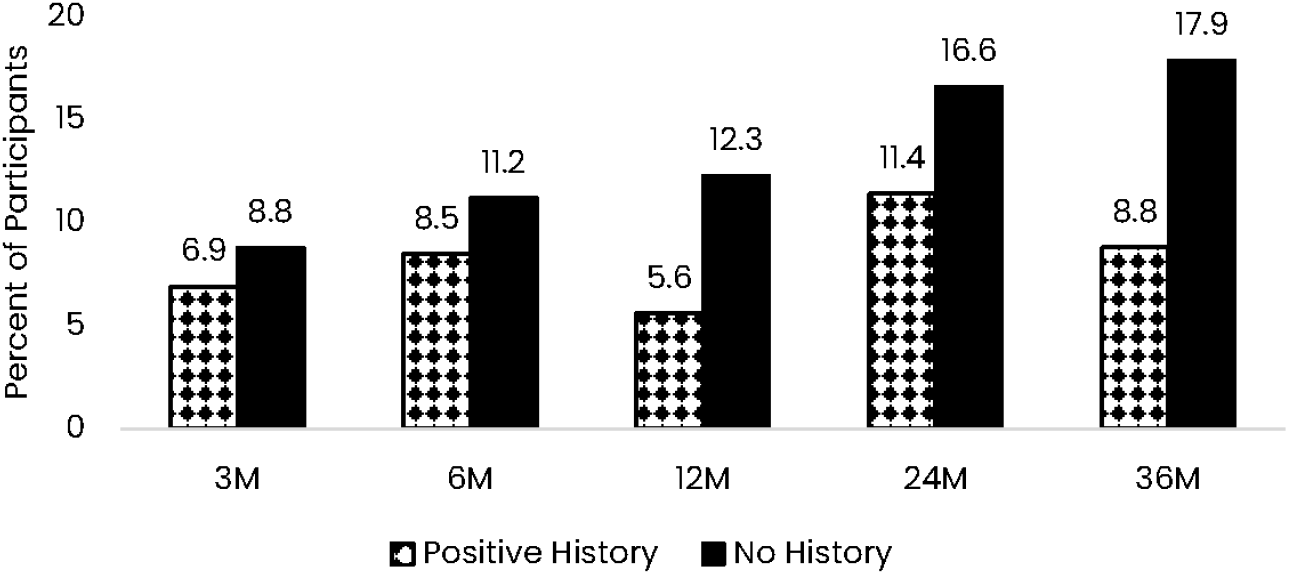
Percent of participants reporting complete seizure freedom at each visit for the 3 months immediately prior

Patterns were consistent within focal and generalized seizure categories (Figure 1). Among participants who reported focal seizures at baseline, median reductions at 36 months were 83.3% (95% CI = 71.88 to 90.91) for those with no previous ICSE and 71.8% (95% CI = 37.27 to 87.50) for those with previous ICSE. Among participants reporting generalized seizures at baseline, the median reduction at 36 months post-VNS implantation was 77.8% (95% CI = 60.17 to 96.88) for no previous ICSE and 76.2% (95% CI = 0 to 95.83) for previous ICSE.

Median percentage reductions from baseline at 36 months between participants with and without prior ICSE across seizure types were not significantly different (p=0.3485, Wilcoxon rank-sum test comparing prior surgery vs. no prior surgery subgroups) (Figure 1). In addition, there were no significant differences across all seizure frequency reduction comparisons (p>0.05 for all time points and seizure types) (Figure 1).

### Adverse events

The overall incidence of adverse events was similar between groups (36.9% vs. 41.2%; IRR = 0.91, 95% CI: 0.70–1.19) (Table 4). Events related to the implant or procedure occurred less frequently in the prior ICSE group (9.5% vs. 13.6%; IRR = 0.65, 95% CI: 0.34–1.26). Rates of events related to stimulation or device were nearly identical (32.1% vs. 32.0%; IRR = 1.04, 95% CI: 0.79–1.38).

**Table 4.** Summary of Incidence Rate (IR) and Incidence Rate Ratio (IRR) by System Organ Class and Preferred Term.

| ***Event Category*** | ***Prior Brain Surgery***<br>***(N=84)*** | ***No Prior Brain Surgery***<br>***(N=447)*** | ***IRR*** | ***95% CI*** |
| --- | --- | --- | --- | --- |
| ***Adverse Events*** | 31 (36.9%)<br>[26.6%, 48.1%]<br>IR=0.27 | 184 (41.2%)<br>[36.4%, 45.8%]<br>IR=0.30 | 0.91 | (0.70, 1.19) |
| ***Related to Implant/Procedure*** | 8 (9.5%)<br>[4.2%, 17.9%]<br>IR=0.04 | 61 (13.6%)<br>[10.6%, 17.2%]<br>IR=0.07 | 0.65 | (0.34, 1.26) |
| ***Related to Stimulation/Device*** | 27 (32.1%)<br>[22.4%, 43.2%]<br>IR=0.25 | 143 (32.0%)<br>[27.5%, 36.4%]<br>IR=0.24 | 1.04 | (0.79, 1.38) |
| ***Serious Adverse Events*** | 3 (3.6%)<br>[0.7%, 10.1%]<br>IR=0.02 | 32 (7.2%)<br>[5.0%, 10.0%]<br>IR=0.03 | 0.56 | (0.20, 1.56) |
| ***Serious AE related to***<br>***Implant/Procedure*** | 2 (2.4%)<br>[0.3%, 8.3%]<br>0.01 | 11 (2.5%)<br>[1.2%, 4.4%]<br>IR=0.01 | 0.69 | (0.16, 3.00) |
| ***Serious AE related to***<br>***Stimulation/Device*** | 0 (0.0%)<br>[0.0%, 4.3%]<br>IR=0.00 | 6 (1.3%)<br>[0.5%, 2.9%]<br>IR=0.01 | 0.32 | (0.02, 5.60) |

Serious adverse events were observed in 3.6% of participants with prior surgery compared to 7.2% without prior surgery (IRR = 0.56, 95% CI: 0.20–1.56), indicating a lower rate in the prior ICSE, though the confidence interval was wide. Serious events related to the implant or procedure were rare in both groups (2.4% vs. 2.5%; IRR = 0.69, 95% CI: 0.16–3.00). No serious events related to stimulation or device occurred in the prior surgery group, compared to 1.3% in the no prior surgery group (IRR = 0.32, 95% CI: 0.02, 5.59).

## Discussion

In this real-world analysis, ICSE did not influence the safety or effectiveness of adjunctive VNS. Seizure frequency reduction at 36 months was comparable between participants with and without prior ICSE, reinforcing the clinical utility of VNS when ICSE has not been effective. Compared with those without prior ICSE, individuals with prior ICSE had similar median reductions in seizure frequency across seizure types and time since implant, but with wider confidence intervals owing to smaller sample sizes. Safety outcomes were similarly consistent across groups. Rates of adverse events, including those related to the implant, procedure, or stimulation, were comparable, and serious adverse events were infrequent in both cohorts. Importantly, no serious stimulation-related events occurred in the prior ICSE group.

The history of prior ICSE was associated with greater variability in seizure outcomes. There was a difference between the two groups in the proportion of individuals achieving a 100% reduction in the number of seizures reported at baseline, with the higher rate in the no prior ICSE group. This difference was almost exactly reversed for the percentage of individuals achieving between 80 and 100% seizure reduction. Several factors may explain this difference. First, patients who undergo ICSE and subsequently require VNS are likely to represent a more refractory subgroup. Surgical intervention is typically reserved for individuals with well-localized epileptogenic zones and carries a high success rate; therefore, those who fail surgery may have multifocal epilepsy (a record of which was not reported in the CORE-VNS case report forms) or more extensive epileptogenic zone/other causes for ongoing seizures. This may reduce the likelihood of complete seizure freedom with any subsequent therapy. Second, people with prior ICSE had a median seizure frequency of 24 seizures per month at baseline, while the group without prior ICSE had a median seizure frequency of 15 seizures per month at baseline, supporting a greater severity of presentation in those having ongoing seizures post-ICSE. Our findings, however, support that PwE who fail to achieve seizure amelioration with ICSE should still be considered for VNS, given the beneficial outcomes noted in this analysis.

Limitations of this analysis include several factors. First, the CORE-VNS study is a prospective, open-label, and observational study. Treatment allocation for this subset analysis, based on ICSE history (positive or negative), was not randomized. Likewise, concomitant ASMs regimens and VNS Therapy programming parameters were not standardized. As a result, causal inferences about the effect of prior surgery on subsequent VNS Therapy cannot be established, and differences between groups may be affected by baseline clinical differences. Second, since this analysis is a post-hoc, the study was not powered to detect minor differences between groups or types of prior ICSE and without consideration for sources of variability. For example, PwE and ICSE in this analysis appear to represent a more refractory population with a higher baseline seizure frequency. In addition, this population was heterogeneous considering the different types of prior surgical procedures (e.g., lesionectomy, lobar resection, callosotomy), timing since surgery, and underlying epilepsy etiology. Finally, as with most long-term real-world studies, incomplete follow-up and missing data may have introduced attrition bias. Despite these limitations, the large, prospective, multinational nature of CORE-VNS provides valuable real-world evidence addressing an important clinical question that has not been systematically evaluated in prior studies.

## Conclusions

This 36-month analysis of the international, multicenter, prospective observational CORE-VNS study showed that the history of ICSE was not associated with changes in seizure outcomes or adverse events in people receiving subsequent VNS therapy. Individuals with a history of prior ICSE showed similar reductions in seizure frequency and adverse event profiles as those without prior ICSE.

## Figure Legends

Figure 1: Median Monthly Seizure Frequency Change and Confidence Intervals by Study Visit. Solid lines represented median change from baseline seizures. Dashed lines represent median 95% CI for each identical colored group.

Figure 2: Percent of participants reporting complete seizure freedom at each visit for the 3 months immediately.

## Authors’ contribution statement

All authors contributed to the conceptualization of the manuscript, critically revised the drafts, and approved the final version for publication. EN was responsible for the conceptualization and writing of the manuscript, compilation of data for figures and tables, critically reviewed and revised drafts, and approved the final version of the manuscript for publication.

## Acknowledgments and funding statement

LivaNova provided financial support for this work. The authors thank Shivani K Maffi for providing editorial support for the submission of the manuscript, which was funded by LivaNova in accordance with GPP3 guidelines.

## Competing interests’ statement

The authors declare the following financial interests/personal relationships which may be considered as potential competing interests: ENN, RV, GG, CS, AM, DM, and RP are employees of LivaNova PLC or a subsidiary, and hold stock or stock options.

KS reports no conflicts of interest related to this publication. There have been previous speaker fees, research grants, and travel reimbursements from Angelini Pharma.

OS, MPF, SB, KR, and NK report no conflicts of interest.

AS is a member of the LivaNova Scientific and Medical Advisory Council. The Oxford Epilepsy Research Group have received speaker fees, travel reimbursement, and consultancy fees from Angelini Pharma, Biohaven, Eisai, LivaNova, Neuro Event Labs, UCB Pharma and Xenon.

FB is an employee of Valos Srl, Genoa, Italy, a partner organization of LivaNova PLC.

## Data availability statement

The data that support the findings of this study are available on request from the corresponding author. The data are not publicly available due to privacy or ethical restrictions.

## Notes

### Clinical Trial

NCT03529045

### Clinical Protocols

https://clinicaltrials.gov/study/NCT03529045

### Author Declarations

The CORE-VNS study involves human participants and was approved by Research Support Services-HREC/18/MonH/198, Ethikkommission des Landes Oberosterreich-1170/2018, Comite d'Ethique Hospitalo- Facultaire- B670201836036, Comissao Nacional de Etica Em Pesquisa-3.583.646, CEP Instituto Estadual do Cerebro Paulo Niemeyer Iecerebro-3849288, CER CHUM- 19.016, MUHC REB-MEO- 02- 2020- 5498, The Western University Health Science Research Ethics Board-113838 and 114135, Renji Hospital Ethics Committee- (Number unknown), CEIm- E-PS2019046, CEIm Provincial de Sevilla-16/2019, Tampere University Hospital Municipality's Local EC-R19007, London- Harrow Research Ethics Committee-18/LO/0552, Institutional Ethics Committees (India)- IEC/1/226/2019, IEC/1254/AL/19/07 and CT_2019_JUL_NK_602, Comitato Etico di Area Vasta Emilia-110/2019/DISP/AUSLBO, Comitato Etico-1353, Ethics Committee in NCNP-A2019- 033, National Hospital Organization Nagasaki Ethics Committee-2019031, METC Isala Zwole-180330, Medisch Ethische Toetsings Commissie-METC 18.20 Medisch Ethische Toetsings Commissie, Erasmus MC- MEC- 2019- 0752, Ethics Committee at Children's Memorial Health Institute-5/ KBE/2019, Comissao de Etica do Centro Hospitalar de Sao Joao-P32- 19, IRB Research Center Jeddah-RC- J/10/41EC, King Faisal Specialist Hospital and Research Center-C380/647/40, Western IRB-20182059, Salus IRB-LNN- 801, Wake Forest Health Sciences IRB-IRB00053820, UTHSC IRB-18- 06378- XP, Ascension Wisconsin IRB-1227, Tulane University Human Research Protection Office-2019- 1419, University of Pennsylvania IRB-832238, UT Health San Antonio IRB-HSC20180684H, Georgetown University IRB-2018- 0874. Participants gave informed consent to participate in the study before taking part. The clinical investigation was performed in accordance with the ethical principles that have their origin in the Declaration of Helsinki, and that are consistent with Good Clinical Practice described in ISO 14155:2011 and ISO14155:2020 guidelines, and the applicable local, regional and national regulatory requirements (including 21 CFR Part 56: Institutional Review Boards). ClinicalTrials.gov ID NCT03529045

